# Vascular Territory, White Matter Connections and the Neuroanatomical Basis of Memory Impairment after Stroke

**DOI:** 10.1101/19012385

**Authors:** Paul Wright, Michael J. O’Sullivan

**Author notes:** **Corresponding author** Professor Michael O’Sullivan, UQ Centre for Clinical Research, Building 71/918, Royal Brisbane and Women’s Hospital Campus, Herston, QLD 4029, Facebook: @ioppn, Twitter: @cosmoslab, @kingsioppn.

## Abstract

**Objective:** To investigate neuroanatomical correlates of memory impairment 30-90 days after stroke, including the role of white matter connections in the core circuit for episodic memory.

**Methods:** A cohort of 179 patients with first symptomatic ischaemic stroke were enrolled into a longitudinal cognitive study, STRATEGIC. Verbal and visual memory were assessed at 50±19 (range 22-109) days. Lesion topography was defined by imaging (n=152). In a representative subgroup (n=53), 3T MRI and tractography was used to define patterns of tract injury and microstructure of uninjured tracts.

**Results:** Lesion location, defined by arterial territory, was associated with verbal memory impairment (F(12,164)=2.62, p=0.003), independent of other factors such as age, risk factor status and lesion volume. Independent lesion symptom mapping identified regions of the left posterior temporal white matter, within the left posterior cerebral artery territory, associated specifically with verbal recall. Visual recognition memory was associated with microstructure of the uninjured fornix but not with lesion location.

**Conclusions:** Infarct location strongly influenced verbal recall performance 50 days after stroke. Damage in two locations underpinned this relationship: the thalamus; and within the left PCA territory, where disconnection of parahippocampal white matter projections contributed to verbal memory impairment in some cases. The correlates of early cognitive prognosis were domain-specific with a different pattern of associations with executive function. The association between microstructure of the uninjured fornix and visual memory might reflect the effects of comorbid pathology on hippocampal circuits.

## BACKGROUND

Memory difficulty is a common sequel of stroke. 43% of stroke survivors in the community report memory difficulties^1^ and objective deficits are confirmed in approximately a quarter of patients by neuropsychological tests^2^. Memory dysfunction is an unmet need^1^ and, in a proportion of patients, goes on to become a cornerstone of post-stroke dementia^3^.

The importance of lesion location in post-stroke memory impairment is incompletely understood. Investigation in large cohorts has often been limited to crude localisation, classifying lesions by hemisphere or cerebral lobes^2^. Conversely, descriptions of strategic locations are derived highly selected case series, so that the importance of these strategic infarct syndromes on a more representative scale is not clear.

In the healthy brain, verbal episodic memory is supported by the extended hippocampal network, which includes anterior thalamic nuclei, mammillary bodies and also key connections between these structures^4, 5^. The fornix is the major route of connection between the hippocampus and diencephalic, basal forebrain and prefrontal regions^5^. In older adults, fornix microstructure is the main correlate of recall performance^6^. Posterior parahippocampal projections also support verbal memory in the normal brain^7^ and become increasingly important in individuals with early memory symptoms^8^. The fornix is rarely affected by ischaemic injury but is highly susceptible to Alzheimer’s disease with structural alterations that are well established by the emergence of first symptoms^8-10^. Parahippocampal connections, in contrast, are often injured by infarction in the middle and posterior cerebral artery territories.

## AIMS

This article is based on analysis of data from the first subacute follow-up time point of STRATEGIC. The objective was to define factors associated with verbal and visual memory after stroke. In a subgroup, diffusion MRI was acquired to determine the extent to which undamaged connections, known to be important to memory in the healthy brain, might contribute to prognosis.

## MATERIALS AND METHODS

### Design and Participants

Patients with first symptomatic ischaemic stroke were recruited within 7 days of onset from a single stroke centre. Inclusion criteria were: age over 50; clinical, supratentorial stroke corroborated by CT or MRI. Exclusion criteria were: previous stroke (clinical or old large-artery infarct on imaging); existing diagnosis of dementia; lack of fluency in English; active malignancy; major neurological or psychiatric disease (defined by DSM-IV-TR); previous moderate to severe head injury (Mayo clinic classification of severity); lack of capacity to consent (including aphasia); other factors precluding cognitive evaluation (e.g. severe visual impairment). Consecutive patients meeting these criteria were invited to take part. Stroke subtype was established based on the Trial of Org 10172 in Acute Stroke (TOAST) criteria, using information from brain imaging and other etiological investigations. All participants gave written, informed consent. The study was approved by the London and Bromley Research Ethics Committee (ref: 13-LO-1745) and is registered with ClinicalTrials.gov (https://clinicaltrials.gov/show/NCT03982147).

### Stroke subtyping, risk factors and infarct delineation

Patients were invited to return for follow-up at a target interval of 30-60 days after stroke (study flowchart, Supplemental Figure I, mean±S.D. interval from onset 49.8±19.3 days, range 22-109 days). Investigation results and risk factor status were reviewed and etiological subtypes of stroke verified or updated. Atrial fibrillation (AF) was defined as >7 mins AF measured by either admission ECG or subsequent prolonged recordings. Carotid stenosis was defined by greater than 50% vessel narrowing on either Doppler ultrasound or CT angiography. Ischaemic heart disease (IHD) was defined by a history of acute coronary syndrome or coronary artery intervention. Smoking and hypertension were defined by self-report. Infarcts involving cortex were classified into anterior (ACA), middle (MCA), or posterior (PCA) cerebral artery territories. The MCA territory was further subdivided into anterior, posterior or deep (striatocapsular territory). Non-thalamic lacunar lesions were defined as lesions in the white matter or deep grey matter with a diameter less than 15mm. Severity of white matter disease on clinical images was graded using the Fazekas scale (see Supplemental Material)^12^.

### Cognitive Evaluation

Six tests was performed at the initial visit: the National Adult Reading Test Revised (NART-R)^13^, to estimate premorbid intellectual function; Montreal Cognitive Assessment (MoCA)^14^; Digit Symbol Substitution Test (DSST), a measure of information processing speed and executive function^15^; Trail-making Test; Digit Span, an index of working memory^15^; and The Free and Cued Selective Reminding Task (FCSRT)^16^. The FCSRT provided the principal evaluation of verbal episodic memory. In this test, participants learn 16 written words presented on 4 cards, each with 4 words belonging to 4 semantic categories, with three rounds or recall, giving a maximum score of 48 for each participant. Additional neuropsychological evaluation of memory was performed in the patients with research MRI (see below), using the Doors and People Test^17^, and a Face Recognition Test of visual memory based on the CAL/PAL Face Database^18,19^.

### Memory-Tractography Substudy

Patients without contraindications to MRI were invited to take part in a research MRI substudy. Scans were performed on a 3T MR750 MR scanner (GE Healthcare) 67.6 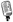 16.5 days after onset(range 30-95 days). T2-weighted fast recovery fast spin echo (FRFSE) and fluid-attenuated inversion recovery (FLAIR) sequences were acquired to delineate infarcts and other vascular lesions (see Supplemental Methods). Lesions were drawn manually by a single rater (PW) using FLAIR images^20^ and tract damage from infarction was inferred from assessing overlap with the JHU white matter atlas.

Diffusion images (see Supplemental Methods for details on acquisition, pre-processing and tractography) were processed as in previous studies^8^. The fornix and parahippocampal cingulum (left and right) were reconstructed with tractography^8^, and used to derive tract-specific measures of microstructure. In two patients with lesions adjacent to the fimbria-fornix on one side, fornix reconstruction was limited to the undamaged hemisphere only.

### Analysis and Statistics

The first analysis tested associations with lesion location by vascular territory (7 regions per hemisphere). Clinical and demographic variables found to have significant univariate associations with cognitive scores (potential confounders for associations between location and cognition) were added as covariates in a univariate General Linear Model for analysis of covariance (ANCOVA). Identical covariates were used to generate comparable models for each of the three cognitive scores.

To corroborate this approach, and to test for anatomical associations at a more fine-grained anatomical level, lesion-symptom mapping was performed using anatomical labels from the AAL and Catani white matter atlases, defining 150 regions of interest per brain. This analysis included 152 of 179 patients with identifiable lesions. Standard space images were analysed using NiiStat (https://www.nitrc.org/projects/niistat/). Differences in verbal recall, DSST and MoCA scores between damaged and intact groups, for each region, were tested using two-sample t-tests, with each score analysed separately. Significance values were corrected using permutation testing with 5000 permutations^21^ (further details in Supplemental Material). Finally, lesion-symptom mapping was also applied to test for associations at the single voxel level.

## RESULTS

### General factors associated with cognition

Verbal recall scores varied significantly by lesion territory (Table 2). Left thalamic and left PCA lesions were both associated with poor verbal recall (Figure 1). An ANOCA model revealed an association between vascular territory and verbal recall that was independent of covariates (F(12,164)=2.62, p=0.003). Lesion vascular territory was not, however, associated with executive function (DSST) or global cognition (MoCA). DSST score was associated with the extent of white matter lesions and diabetes mellitus (Table 2), with DSST scores decreasing monotonically with increasing Fazekas score (Supplementary Figure IV).

**Table 1.**
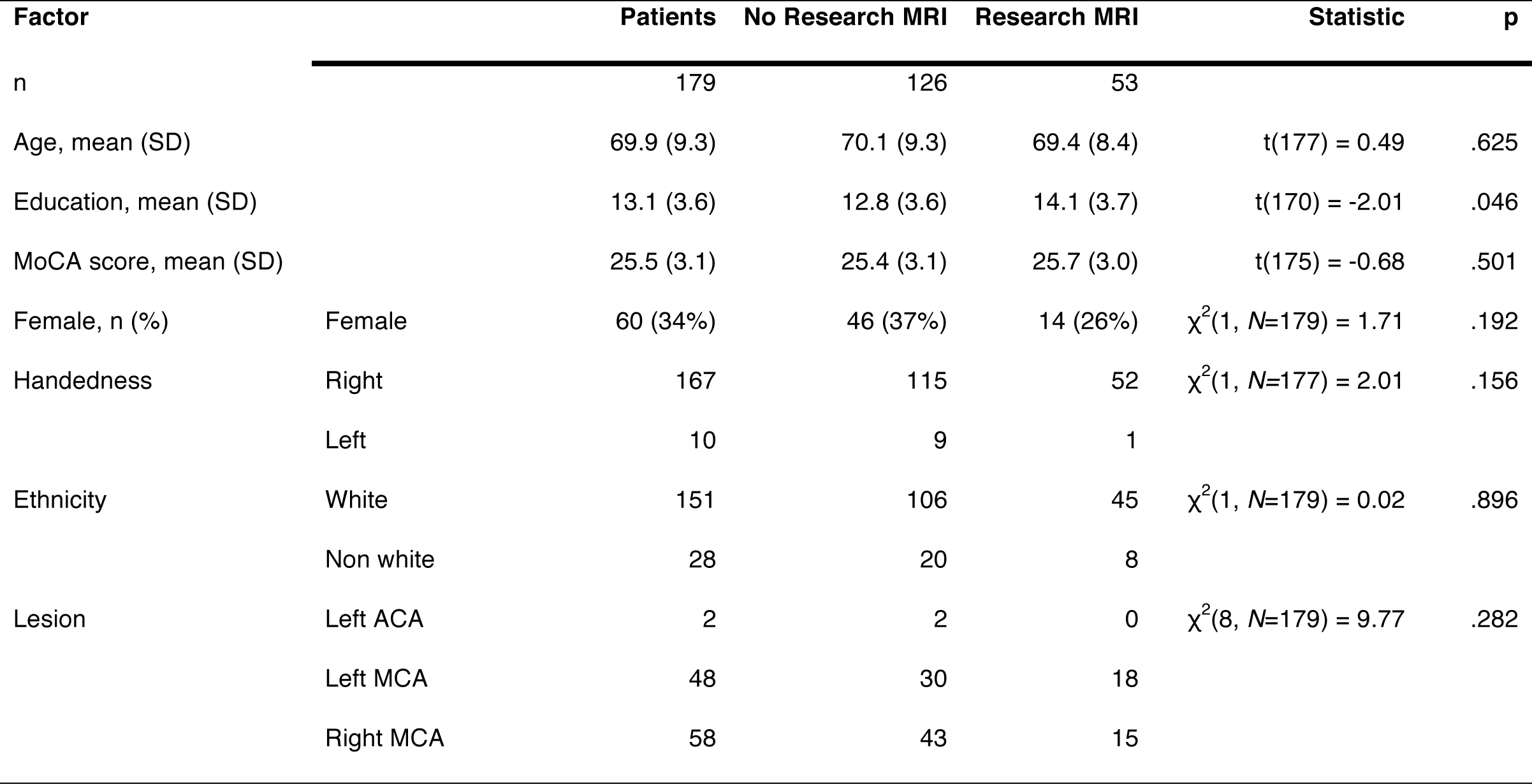
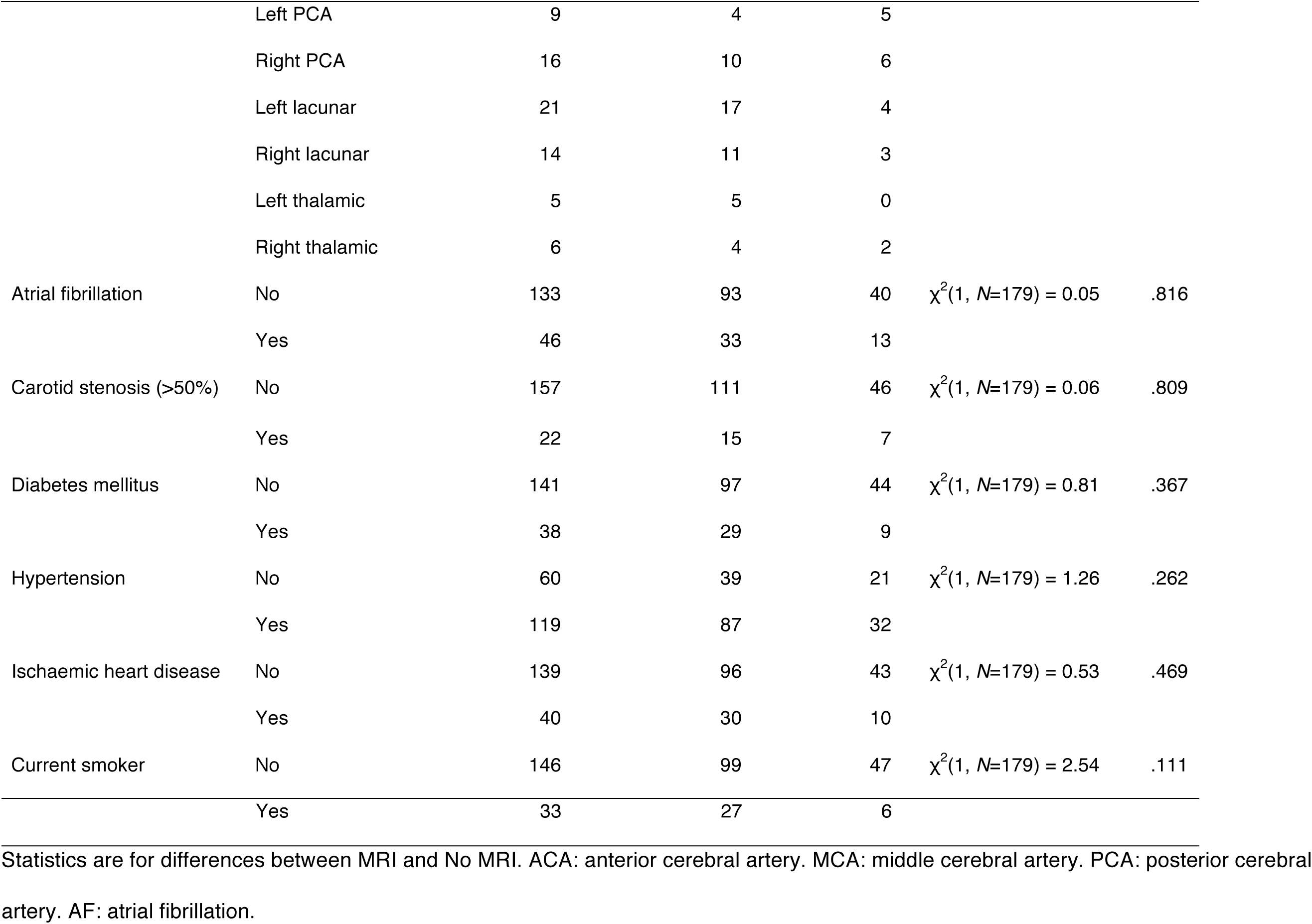
Participant demographics, infarct locations and risk factors.

**Table 2.**
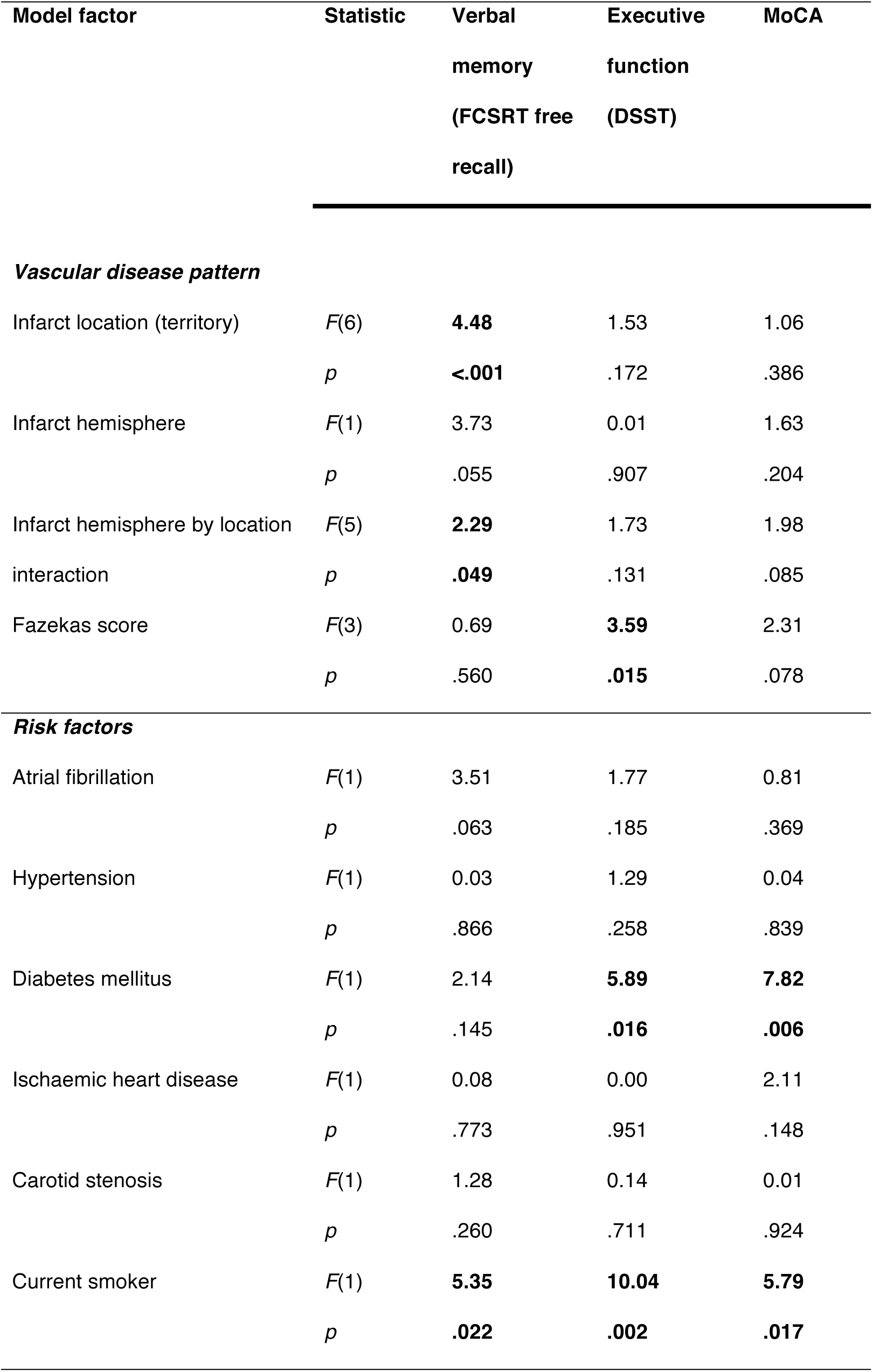
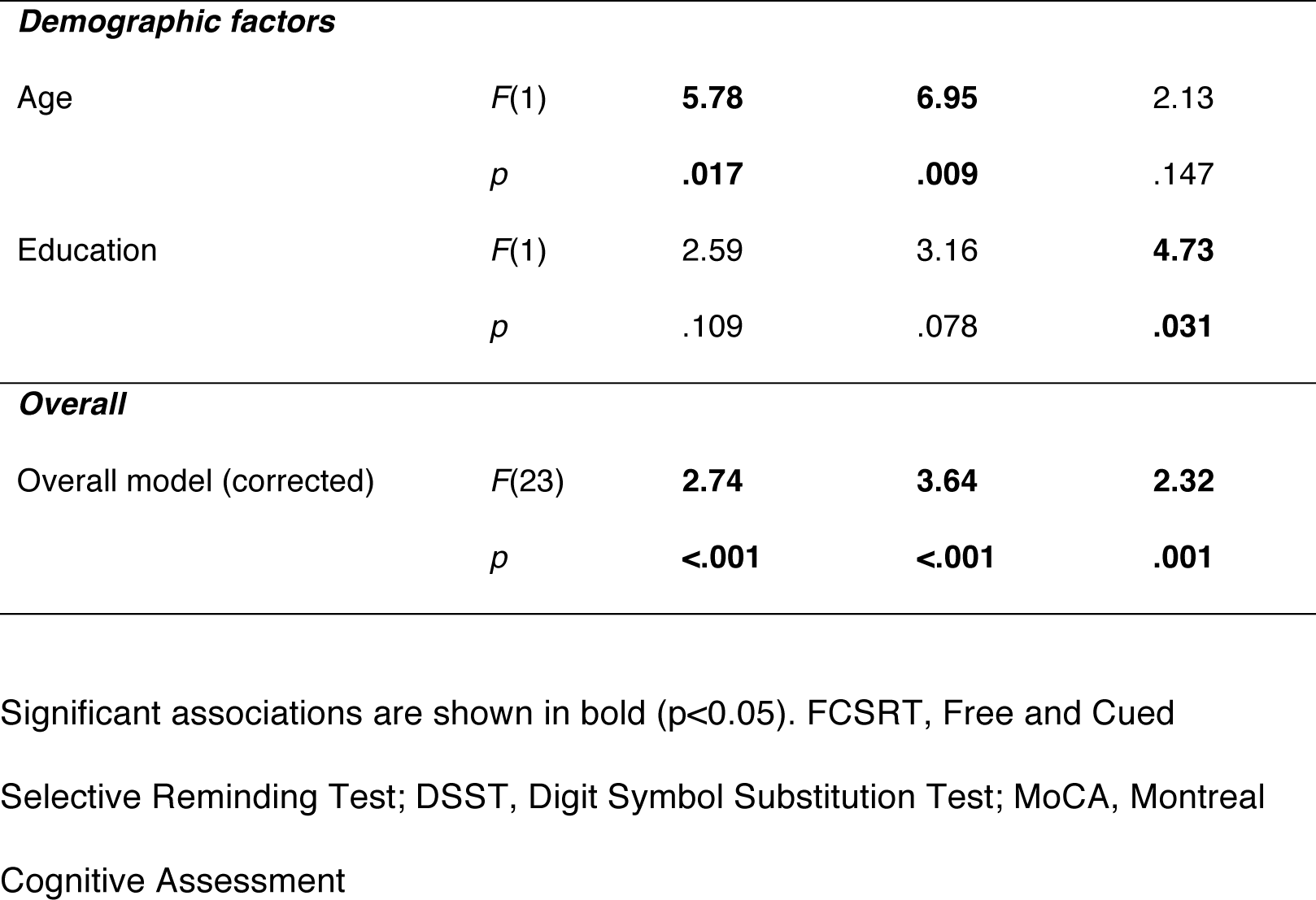
Predictors of memory, executive function and global cognition

| Model factor | Statistic | Verbal<br>memory<br>(FCSRT free<br>recall) | Executive<br>function<br>(DSST) | MoCA |
| --- | --- | --- | --- | --- |
| ***Vascular disease pattern*** |  |  |  |  |
| Infarct location (territory) | *F*(6) | **4.48** | 1.53 | 1.06 |
|  | *p* | **<.001** | .172 | .386 |
| Infarct hemisphere | *F*(1) | 3.73 | 0.01 | 1.63 |
|  | *p* | .055 | .907 | .204 |
| Infarct hemisphere by location<br>interaction | *F*(5) | **2.29** | 1.73 | 1.98 |
|  | *p* | **.049** | .131 | .085 |
| Fazekas score | *F*(3) | 0.69 | **3.59** | 2.31 |
|  | *p* | .560 | **.015** | .078 |
| ***Risk factors*** |  |  |  |  |
| Atrial fibrillation | *F*(1) | 3.51 | 1.77 | 0.81 |
|  | *p* | .063 | .185 | .369 |
| Hypertension | *F*(1) | 0.03 | 1.29 | 0.04 |
|  | *p* | .866 | .258 | .839 |
| Diabetes mellitus | *F*(1) | 2.14 | **5.89** | **7.82** |
|  | *p* | .145 | **.016** | **.006** |
| Ischaemic heart disease | *F*(1) | 0.08 | 0.00 | 2.11 |
|  | *p* | .773 | .951 | .148 |
| Carotid stenosis | *F*(1) | 1.28 | 0.14 | 0.01 |
|  | *p* | .260 | .711 | .924 |
| Current smoker | *F*(1) | **5.35** | **10.04** | **5.79** |
|  | *p* | **.022** | **.002** | **.017** |

| **Demographic factors** |  |  |  |  |
| --- | --- | --- | --- | --- |
| Age | *F*(1) | **5.78** | **6.95** | 2.13 |
|  | *p* | **.017** | **.009** | .147 |
| Education | *F*(1) | 2.59 | 3.16 | **4.73** |
|  | *p* | .109 | .078 | **.031** |
| **Overall** |  |  |  |  |
| Overall model (corrected) | *F*(23) | **2.74** | **3.64** | **2.32** |
|  | *p* | **<.001** | **<.001** | **.001** |

Significant associations are shown in bold ( $p < 0.05$ ). FCSRT, Free and Cued Selective Reminding Test; DSST, Digit Symbol Substitution Test; MoCA, Montreal Cognitive Assessment

**Figure 1.**
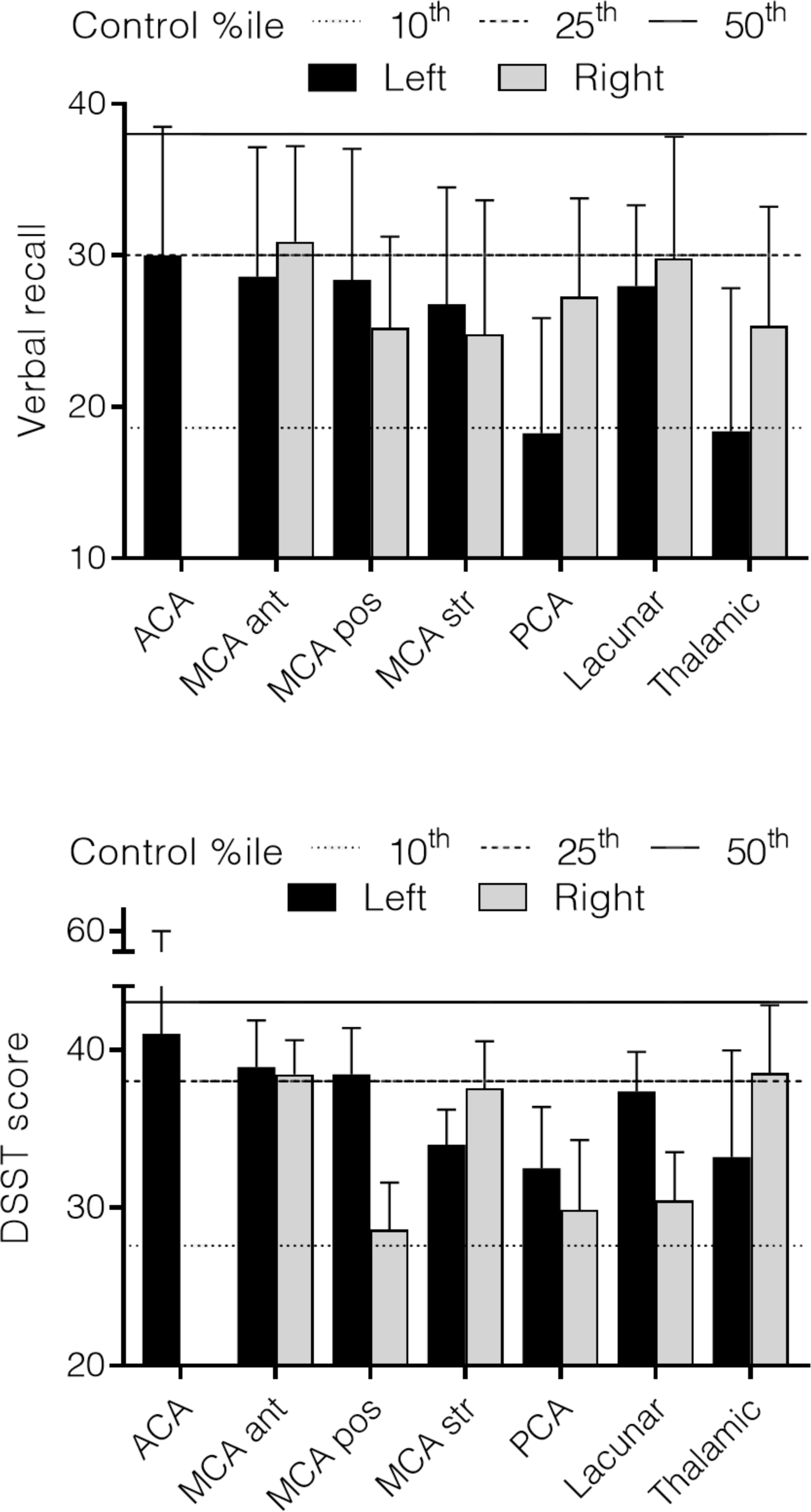
Lesion location and episodic memory impairment 30-90 days after stroke. Mean verbal free recall scores for patients grouped by infarct hemisphere and territory. Left PCA and left thalamic groups fall below the 5% percentile score for healthy participants (dashed line). ACA: anterior cerebral artery. MCA ant: middle cerebral artery, anterior. MCA pos: middle cerebral artery, posterior. MCA str: middle cerebral artery, striatocapsular. PCA: posterior cerebral artery. Bars indicate one standard error of the mean.

### Regional Lesion-symptom mapping

Sites of injury associated with verbal recall were limited to the left hemisphere. Four regions met the permutation-corrected threshold for significance: the left hippocampus (z = -3.24), the left thalamus (z = -3.75), the left cingulum (z = -3.32) and the left inferior occipito-frontal fasciculus (z = -3.54). Executive function was associated with damage in the right hippocampus (z = -3.25) right parahippocampal gyrus (z = -3.49) and right cingulum (z = -3.76). Voxel-level analysis revealed a similar pattern with only a cluster in the left thalamus reaching whole brain significance.

### Uninjured Connections and Memory: the Fornix

Visual recognition memory scores correlated with mean diffusivity of the unaffected fornix (face recognition total hits, r = -.38, p < .01; remembered hits r = -.45, p < .001; Doors and People verbal recognition r = -.30, p < .05; visual recognition r = -.23, p = .058). Fornix microstructure was not associated with verbal recall, creating a double dissociation for verbal and visual memory (Figure 3).

**Figure 2.**
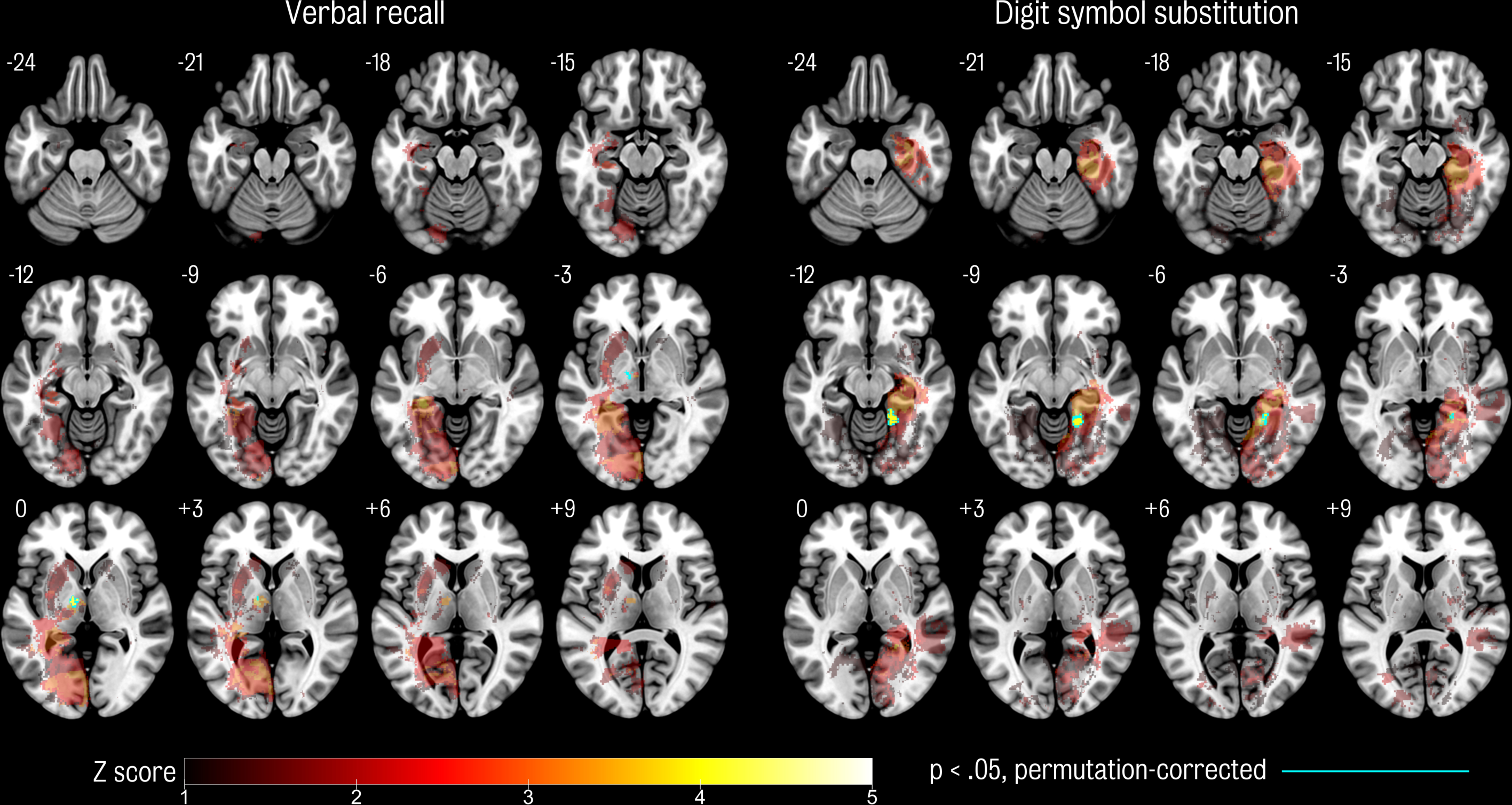
Voxel-based lesion-symptom mapping. Colour maps show the significance of the association between voxel lesion status and cognitive score. Results are shown at an uncorrected threshold of Z = 2.33 (approximately p < .01). Effects surviving permutation correction are outlined in blue. Numbers to the top left of each brain section indicate the MNI coordinate.

**Figure 3.**
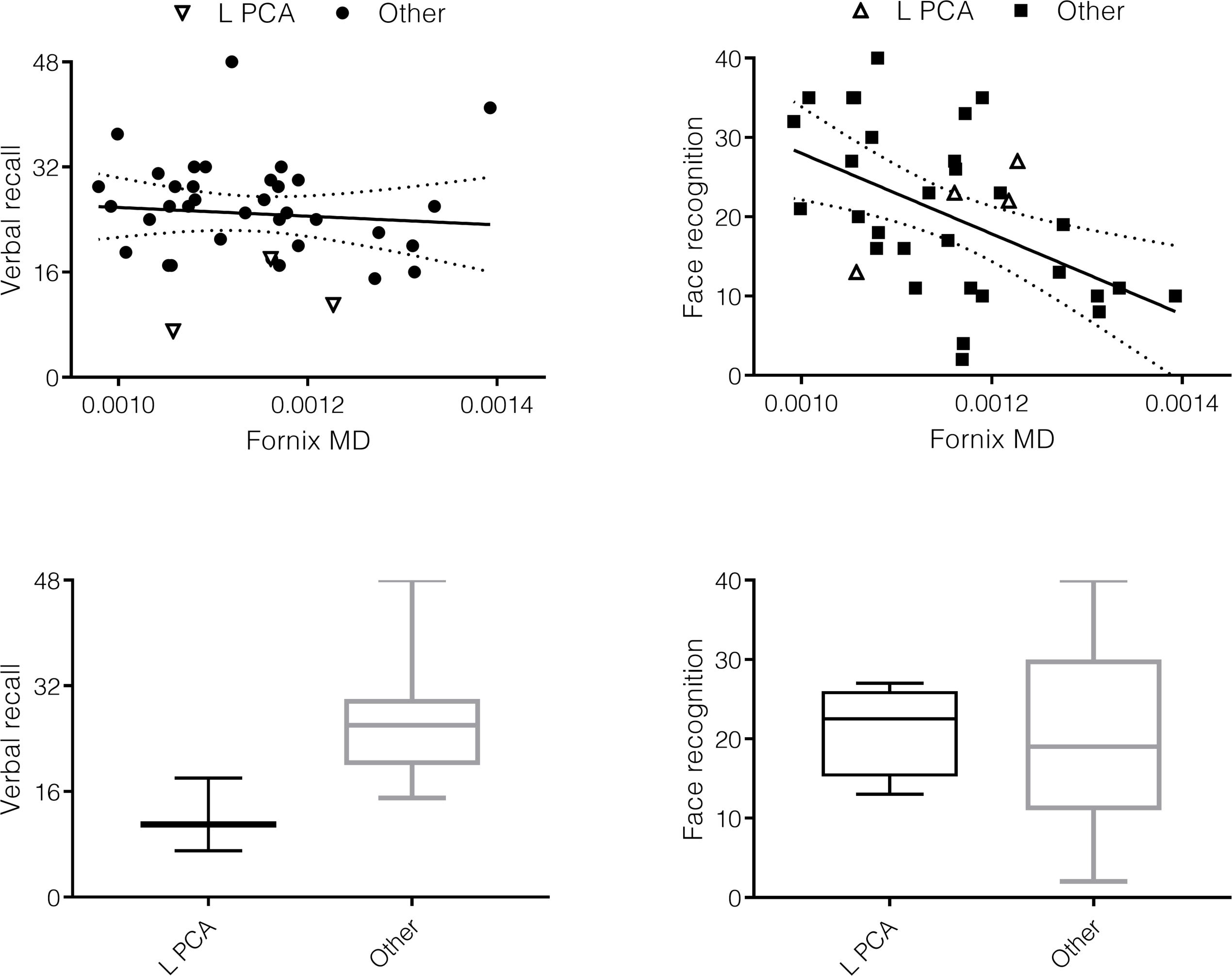
Dissociable relationships between: i.) verbal recall memory and lesion location; ii.) visual recognition memory and fornix microstructure. Patients show dissociated relationships between verbal recall and lesion type (left column) and between face recognition and fornix microstructure (right column). Scatterplots (top row) show the relationship between fornix MD and face recognition remembered hits (r = -.43, p < .01, controlling for age), but patients with strategic lesions (unfilled triangles) are interspersed with patients with other lesions. In contrast, verbal recall has no relationship with fornix MD, but patients with strategic lesions score lower than the other group (t(36) = 3.30, p < .01; see boxplots in bottom row).

## DISCUSSION

In a sample of 179 patients with first symptomatic ischaemic stroke, lesion location was found to have the strongest influence on verbal recall performance in the first three months after stroke. Lesion location, represented by vascular territory, remained a significant factor after accounting for other variables in a multi-variable model. In contrast, vascular territory was not associated with executive function (DSST) or MoCA score. An independent lesion-symptom mapping analysis evaluated 150 regions in the brain and identified critical white matter regions within the left PCA territory. Age, smoking and infarct volume were associated with cognitive function in univariate analyses but these associations were not sustained once other variables were taken into account, which is consistent with previous investigations^2^.

The role of posterior temporal white matter pathways in memory has been a topic of controversy^23^. One previous case series suggested that unilateral posterior temporal infarction was sufficient to cause amnesia^24^. However, this series was based on 1980s CT imaging only, which provides limited sensitivity to hippocampal injury. A subsequent study of experimental PCA lesions in the macaque suggested that memory was only impaired when the hippocampus was injured^25^. The present analysis was able to exclude involvement of the hippocampus in most cases and reasserts the message that damage to posterior temporal white matter can cause marked verbal memory impairment in the absence of overt hippocampal injury^24^. The most likely substrate – supported by analysis of a number of individual cases in this cohort (Figure 4) – is damage to the parahippocampal portion of the left cingulum bundle (left PHC) which has been found to have a role in verbal memory in healthy volunteers^8^. Interestingly, the left PHC seems to have a disproportionate importance for supporting memory in individuals who have already developed mild cognitive impairment^8^. Memory deficits in patients with left PCA lesions were often unrecognised clinically, perhaps because the hippocampus was not obviously injured. This illustrates the importance of awareness of the risk of memory dysfunction in patients with left PCA infarction.

**Figure 4.**
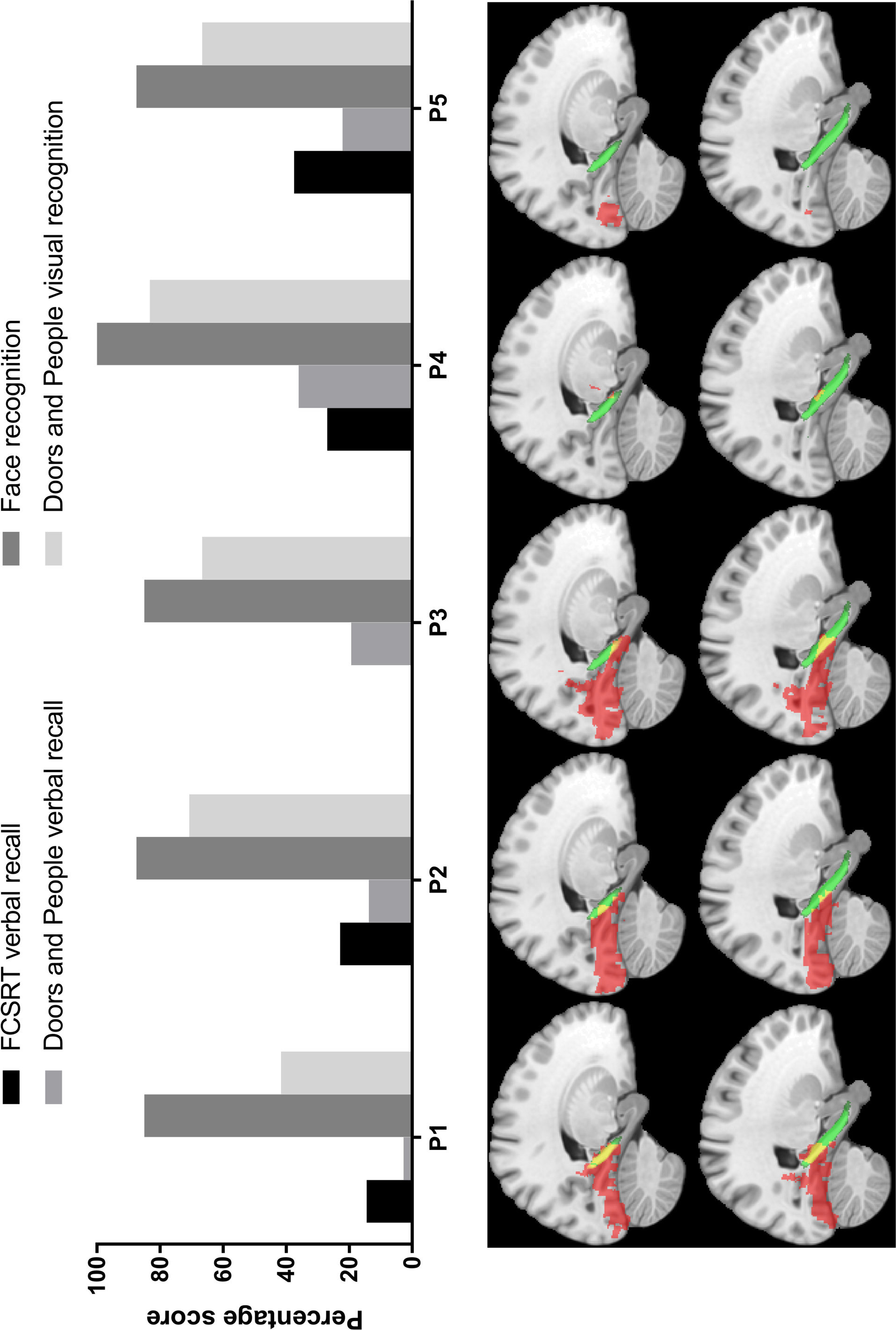
Disconnection of the left parahippocampal connections in individual cases of left PCA stroke. Top: score for each of the five left PCA patients on tests of verbal recall and visual recognition. Bottom: sagittal sections of a standard MNI brain template (co-ordinates x = -20 and x= -24 mm). Each infarct is shown in red; the left parahippocampal cingulum (PHC), defined by the JHU atlas is shown in green; the overlap in yellow. Verbal recall scores were lowest in the individual with most marked overlap of the PHC (far left). FCSRT: Free and Cued Selective Reminding Test. (P3 was unable to perform FCSRT verbal recall).

At a single voxel-level only a cluster of voxels within the left thalamus retained significant association with verbal recall. Executive function was associated with a distinct set of regions in the right hemisphere. Although the study lacked the power to infer relationships with cognition at a single-voxel level, it still represents an approximate 20-fold improvement in the anatomical precision of associations between lesion location and cognition, which in previous literature was limited to location by hemisphere or lobe of the brain, in contrast to a parcellation of the brain into 150 separate regions in the regional lesion-symptom mapping analysis.

Microstructure of the fornix correlated with visual recognition memory performance independent of lesion volume and location. Measurements of fornix microstructure were restricted to undamaged white matter. These results indicate that both the direct effects of injury and the status of critical connections remote from injury contributed to the profile of memory impairment. One possible interpretation is that secondary degeneration of the fornix, caused by the index infarct, had already developed when imaging took place at 30-90 days. This explanation seems unlikely as fornix microstructure was not associated with volume of infarction or the interval between clinical onset and imaging. An alternative explanation is that the microstructure of the fornix provides a potential biomarker of early neurodegenerative pathology^9, 10^. Population-based studies suggest that undiagnosed mild cognitive impairment may be present in up to 37% in individuals aged 70-90 years^26^.

The current sample was limited to patients who had capacity to consent and who could perform cognitive tests involving reading, writing and verbal responses. Thus, our findings do not extend to patients with more severe strokes or specific deficits such as aphasia or alexia. The current analyses show that approximately 30% of the variance in verbal memory at 30-90 days can be accounted for by the features of the incident infarct, structure of important connections, and background factors such as age and risk factor status. The extent to which secondary or independent structural changes in the cortex, thalamus or striatum – assessed, for example, by cortical thickness measurements and assays of vascular lesions not visible on 3T MRI – are able to account for additional variance in cognitive outcome is an important topic for future analyses. A limitation of lesion symptom mapping approaches is that the power to make inferences about any one region depends on frequency of involvement of that region. In the present study, there were more lesions, and therefore more power, in the right hemisphere. The lateralisation of associations with executive function is likely to reflect this factor. The current study only included patients with a single large artery lesion and therefore cannot elucidate additive or synergistic impairments from multiple lesions. This important issue would require a much larger study sample to address using multivariate statistical methods.

## Data Availability

All data is available for sharing with qualified investigators and will be shared, in anonymized form, upon request, in accordance with the terms of the study ethical approvals.

